# Sex Differences in Colonic Mucosal Microbiome of Irritable Bowel Syndrome Patients Compared to Healthy Controls

**DOI:** 10.1101/2024.07.31.24311317

**Authors:** Swapna Mahurkar-Joshi, Simer Shera, Jennifer Labus, Tien S. Dong, Jonathan P. Jacobs, Lin Chang

## Abstract

**Background:** Irritable bowel syndrome (IBS) is a female-predominant disorder of brain-gut interactions. Our previous study on colonic mucosal microbiota demonstrated significant differences between IBS bowel habit subtypes and showed that gut microbiota is associated with abdominal pain in IBS patients. However, there is no consensus on sex-related differences in mucosal microbiota in IBS compared to healthy controls (HC). We aimed to identify sex-related differences in the mucosal microbes associated with IBS.

**Methods:** Sigmoid mucosal biopsies were obtained from 97 Rome+ IBS patients and 54 healthy controls (HC). Mucosal microbiome was characterized using 16S rRNA sequencing and analyzed and general linear models were used to test group differences between IBS diagnosis and sex. Sex-specific relationships between mucosal microbiome and IBS symptoms were assessed using sparse partial least squares (sPLS) regression.

**Results:** Beta diversity was significantly different between men and women overall (p=.03) but not within IBS or HC. IBS women showed lower abundance of *Catenibacterium* and *Ruminoclstridium_9* and increased abundance of *Bacteroides*, *Escherichia/Shigella, Lachnoclostridium* and *Ruminococcaceae* compared to men with IBS (p<0.05). However, healthy women had a lower abundance of six distinct genera compared to healthy men. In women, higher IBS symptoms were associated with an increased abundance of bacteria including *prevotella_9,* and *paraprevotella*, however, in men, IBS symptoms were associated with increased abundances of genera such as *Dialister*. Interestingly, increased abundance of *Desulfovibrio* was associated with higher symptoms in women but lower symptoms in men.

**Conclusion:** There are distinct sex-related differences in the mucosal microbiome between IBS and healthy participants supporting the importance of studying sex-specific mechanisms in IBS pathophysiology.

## Introduction

Irritable bowel syndrome (IBS) is a female-predominant disorder of altered bidirectional gut-brain interactions characterized by chronic abdominal pain and altered bowel habits.^1, 2^ It is a widely prevalent condition affecting up to 11% of the general population.^3^ IBS is associated with microbial dysbiosis and studies suggest a key role for the gut microbiome in its pathogenesis.^4–7^ Most studies investigating relationships between the gut microbiome and IBS focused on fecal samples due to their noninvasive collection methods.^8^ While these studies have provided valuable insight, fecal samples do not capture critical microbial interactions occurring at the mucosal surface. Emerging evidence suggests that mucosal microbiota, or microbiota in close proximity to the gut epithelium layer, immune cells and enteric nerves, may play a pivotal role in IBS.^9, 10^ Our earlier work identified significant IBS bowel habit differences in mucosal microbiota, such as constipation-predominant IBS (IBS-C), diarrhea-predominant IBS (IBS-D), and IBS with mixed bowel habits (IBS-M), and reported an association between gut microbiota composition and abdominal pain in IBS patients.^11^ These findings emphasize the importance of considering mucosal microbiota when investigating IBS.

The influence of sex on gut microbiome has not been well studied in IBS, despite IBS being more common in women compared to men and women reporting a higher symptom severity.^12–14^ We recently found sex differences in fecal microbial composition in IBS participants but not healthy controls (HC). *Prevotella* was the main difference between men and women with IBS and it negatively correlated with IBS severity^15^. However, sex-specific differences in mucosal microbiota and their association with IBS have not been investigated. Therefore, this study aimed to identify 1) sex differences in mucosal microbial abundances overall and within IBS and HC groups and 2) mucosal microbiota associated with IBS symptom severity in women and men.

## Methods

### Study participants

The study population included 18-55 year-old men and women with IBS and HC recruited by community advertisement as described previously.^11^ IBS and bowel habit subtypes were diagnosed based on medical history, physical exam, and Rome criteria.^2^ Subtypes included IBS-C, IBS-D, IBS-M, and unsubtyped IBS (IBS-U). HC had no personal or family history of any pain conditions. Additional exclusion criteria for all participants included the following: celiac disease, inflammatory bowel disease, or other gastrointestinal (GI) illness which could explain IBS symptoms, active psychiatric illness in the past six months according to the Diagnostic and Statistical Manual of Mental Disorders, 4th Edition, Text Revision (MINI International Neuropsychiatric Interview),^16^ current substance dependence or abuse, smoking more than 1/2 pack per day, and use of corticosteroids in past six months, antibiotics or narcotics in past three months, or probiotics in past one month before the procedure. Participants were not excluded if they were on proton pump inhibitors. During the procedure, two patients were reported using pantoprazole or omeprazole. The University of California, Los Angeles Institutional Review Board approved the study, and participants signed a written informed consent before the study. Participants were compensated.

### Questionnaires

Patients were assessed for overall severity, abdominal pain, and bloating within the prior week with numerical rating scales from 0 (no pain) to 20 (most intense symptoms imaginable) using our Bowel Symptom Questionnaire (BSQ).^17^ The Hospital Anxiety and Depression Scale (HAD) was used to measure current anxiety and depression symptoms.^18^ The IBS-SSS assesses the severity and frequency of abdominal pain, distention, bowel dysfunction, and global well-being in IBS.^19^ Summed scores between 75 and 175 represent mild disease, 175-300 moderate disease, and >300 severe disease. Each questionnaire is validated and widely used, with instructions for how to complete and score them.

### Sample Collection

During flexible sigmoidoscopy, mucosal biopsies were collected at 30 centimeters from the anal verge. Biopsies were snap-frozen in liquid nitrogen and stored at −80C. Participants were asked to withhold from using aspirin and nonsteroidal anti-inflammatory drugs 72 hours before the procedure.

### Dietary Assessments

An institution-developed Diet Checklist was used with the intention to represent diet(s) that best reflect what individuals consume regularly.^20^ Options included standard or modified American, Mediterranean, vegetarian, vegan, dairy-free, gluten-free, and/or low fermentable oligosaccharides, disaccharides, monosaccharides, and polyols (FODMAP) diets. Each diet was classified as either standard or restrictive depending on the elimination of certain food groups by choice. A food frequency questionnaire (FFQ) and diet diary were used for a specific subset of patients to validate the diet checklist.^20^

### Microbiome Analysis

#### DNA Extraction and Sequencing

Bacterial DNA was extracted from the biopsies using the PowerSoil DNA Isolation Kit (MO BIO Laboratories) with bead beating. 16S rRNA gene sequence libraries were generated using the V4 hypervariable region and amplified with the 515F and 806R primers. Libraries were sequenced on Illumina HiSeq 2500 platform (Life Sciences, Branford, CT).

The DADA2 pipeline in R was used to process the raw reads.^20^ Silva v132 reference database was used to eliminate chimeras and assign taxonomy. Samples were rarefied to even depth for calculation of alpha diversity measures using the function, “rarefy_even_depth.” The read depth was 29,709 (SD: 33,526.25) and rarefied to 538 counts. To avoid data loss, alpha diversity was calculated using unrarefied data and beta diversity using relative abundance. Amplicon sequence variant (ASV) data was agglomerated at the genus level to study shifts in microbial abundance. ASVs are exact sequence variants compared with operational taxonomic units, or clusters of reads with 97% similarity.

#### Statistical Analysis

All statistical analyses were performed using R software.^21^ Sex differences in alpha diversity and beta diversity overall and within IBS and HC were assessed using a general linear models. The model included age, body mass index (BMI), race, intake of a restrictive diet, and sequencing batch as covariates. Frequent category imputation for categorical data and mean values for continuous data were used to address missing data on covariates. To maximize power, the restrictive diet variable was imputed for missing data when used as a covariate. Chao1 and Shannon’s index were used as metrics for alpha diversity as described previously^11^. Briefly, beta diversity was evaluated using DEICODE, a form of Aitchison distance that is robust to high levels of sparsity, implemented in a QIIME environment. Adonis, a permutational analysis of variance, was used to test for associations with demographic characteristics and clinical factors.

Group comparisons for clinical and demographic characteristics were performed using t-test or chi-square tests. Genus-level data were extracted and filtered to include genera present in atleast 10% samples. Data were transformed using centered log-ratio (CLR) transformation. Generalized linear (GLM) with sex as an independent variable, microbial abundance as an independent variable covarying for age, BMI, race, intake of restrictive diet, and technical sequencing batch as covariates, was used to evaluate differences in microbial abundances between females and males. We also evaluated sex differences within IBS and HC groups within the framework of a general linear model with microbial abundance dependent variable. Bowel habit was used as a covariate when analyzing group differences within IBS. Results were corrected for multiple comparisons using a false discovery rate (FDR) and an FDR<.25 was considered significant.

Sparse partial least squares (sPLS) regressions were performed to reduce dimensionality while analyzing correlations between clinical variables and microbiota data in multivariate analysis.^22^ Data were residualized for sequencing batch, diet, and bowel habit subtypes. The “tune.spls” function from ‘mixOmics’ package was used to select tuning parameters and the first two components were used for analysis.^23^

## Results

### Demographic and clinical characteristics

We analyzed sex differences in colonic mucosal microbiome of 151 participants (97 Rome+ IBS patients and 54 healthy controls). **Table 1** shows the demographic characteristics of male and female IBS and HC participants. IBS women were younger compared to IBS men (mean [standard deviation] = 29.4 [10.8] vs 36.3 [10.3], p=0.003), and there were differences in the bowel habit subtypes with more men with IBS-D subtype compared to IBS-C (Chi-squared = 14.66, p=0.02). There were no other significant differences in demographic characteristics between sexes within both groups. Additionally, IBS symptom scores were comparable between men and women with IBS (**Table 1**).

**Table 1:**
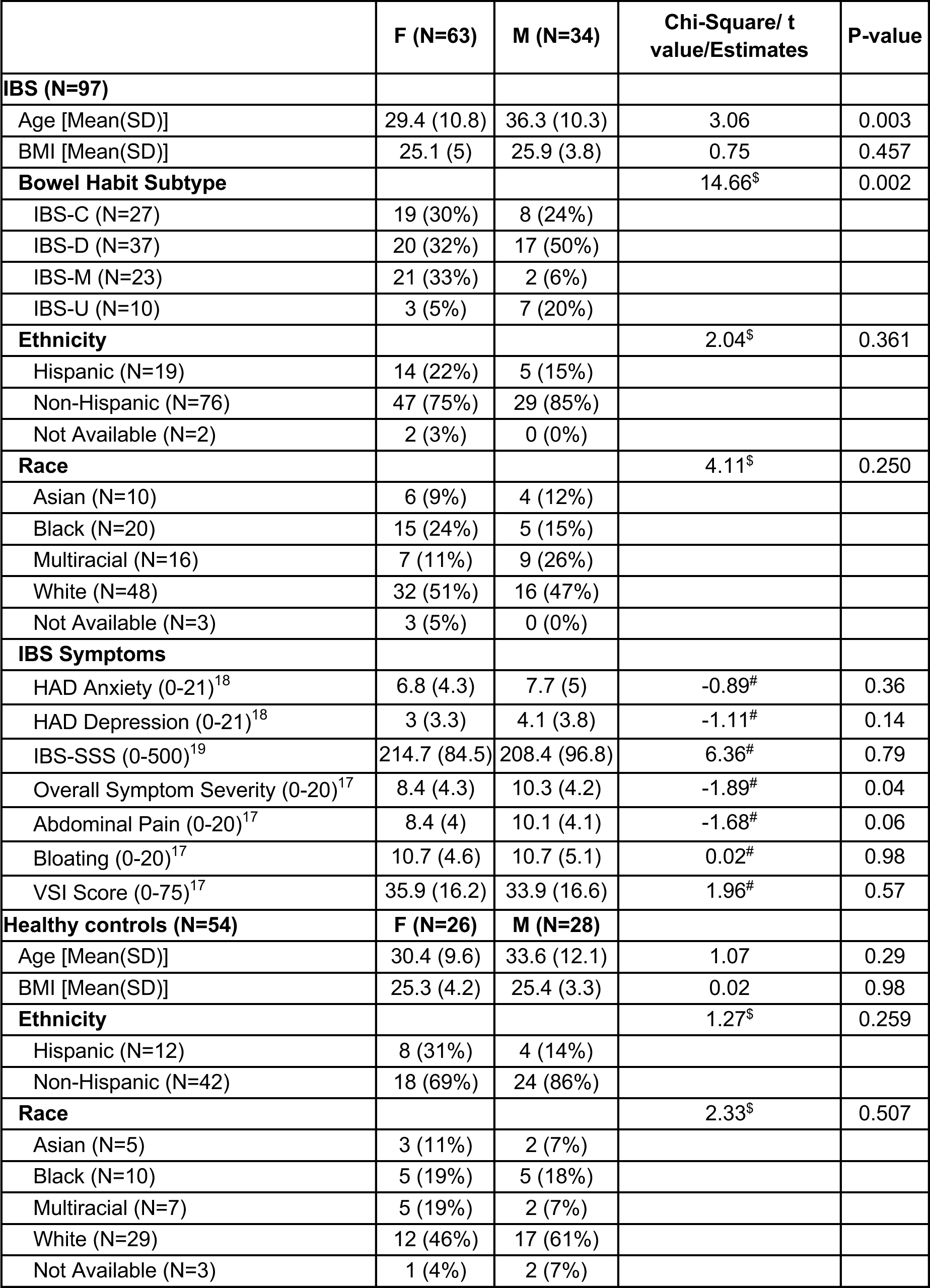
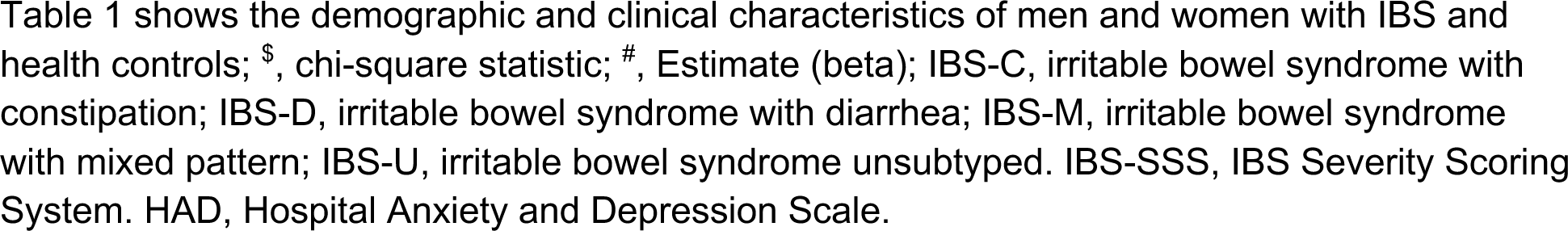
Demographic and Clinical Characteristics of the Study Population.

### Mucosal Microbiota in Men Compared to Women with IBS and Healthy Controls

Microbial analysis revealed significant differences in beta diversity between men and women in the overall study population (r^2^=0.014, p=0.03). There were no significant differences in alpha diversity. Although no genera were significant at an FDR<.25, we found nominal significance (p<0.05) for differences between sexes. Genus-level differences were found in the abundance of five microbes between women and men in the overall participant population. Three genera (*Prevotella_9*, *Catenibacterium, and Ruminiclostridium_9*) were less abundant and two (*Tyzzerella* and *Escherichia/Shigella*) were more abundant in women compared to men (**Table 2**). Within the IBS group, women showed an altered abundance of five genera including a lower abundance of *Catenibacterium,* and *Ruminoclstridium_9*, and an increased abundance of *Bacteroides*, *Escherichia/Shigella*, and *Lachnoclostridium* compared to men (**Figure 1A**, **Table 2**). However, within the HC group, women had a lower abundance of six genera compared to men (**Figure 1B**, **Table 2**). The sex-associated microbes within IBS were distinct compared to HC.

**Figure 1.**
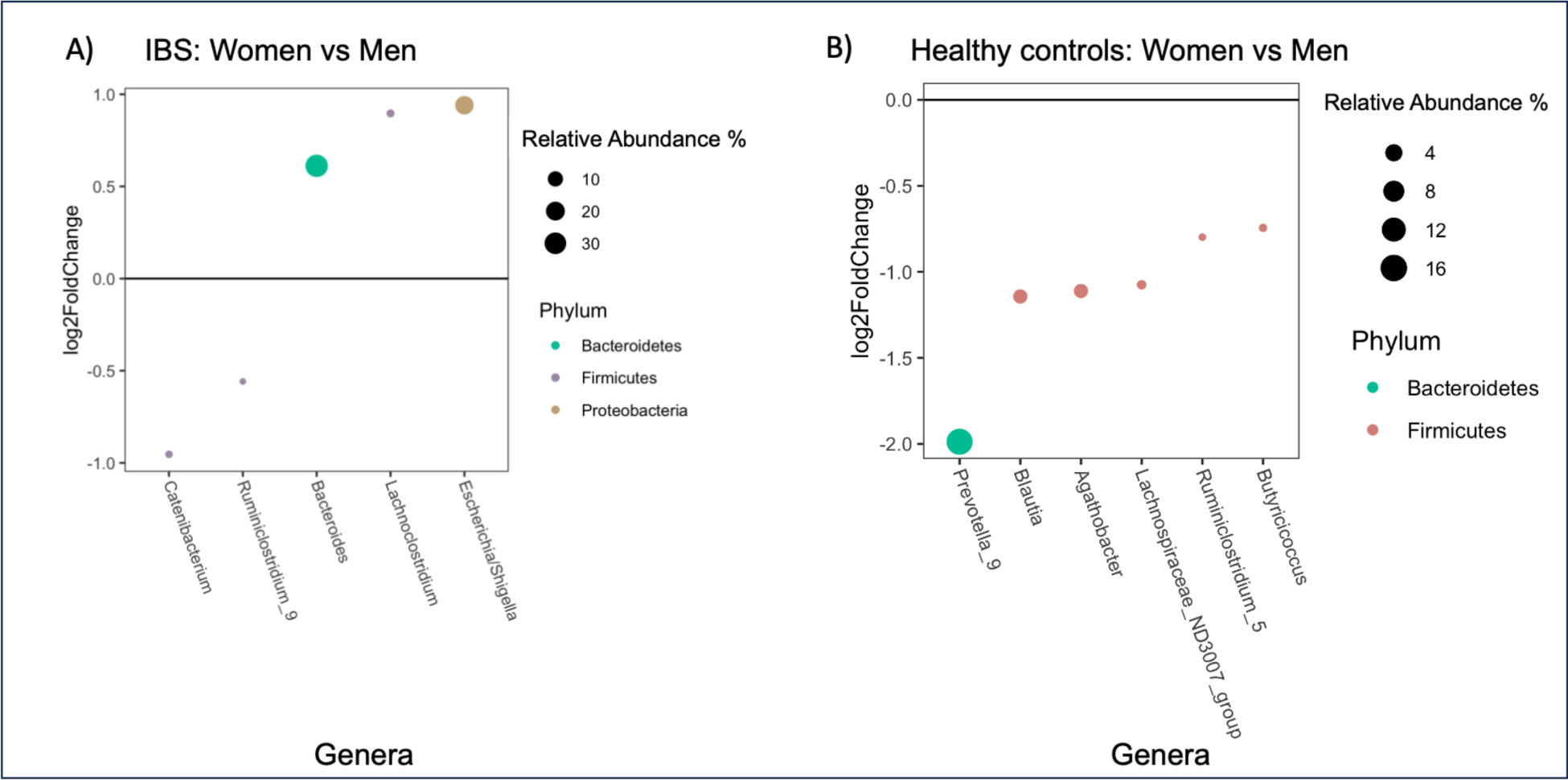
Microbial Genera Altered in Women Compared to Men in IBS Patients and Healthy Controls. At the genus level, five microbes were different between women compared to men in IBS and six microbes were different between women compared to men in healthy controls (p<0.05).

**Table 2:**
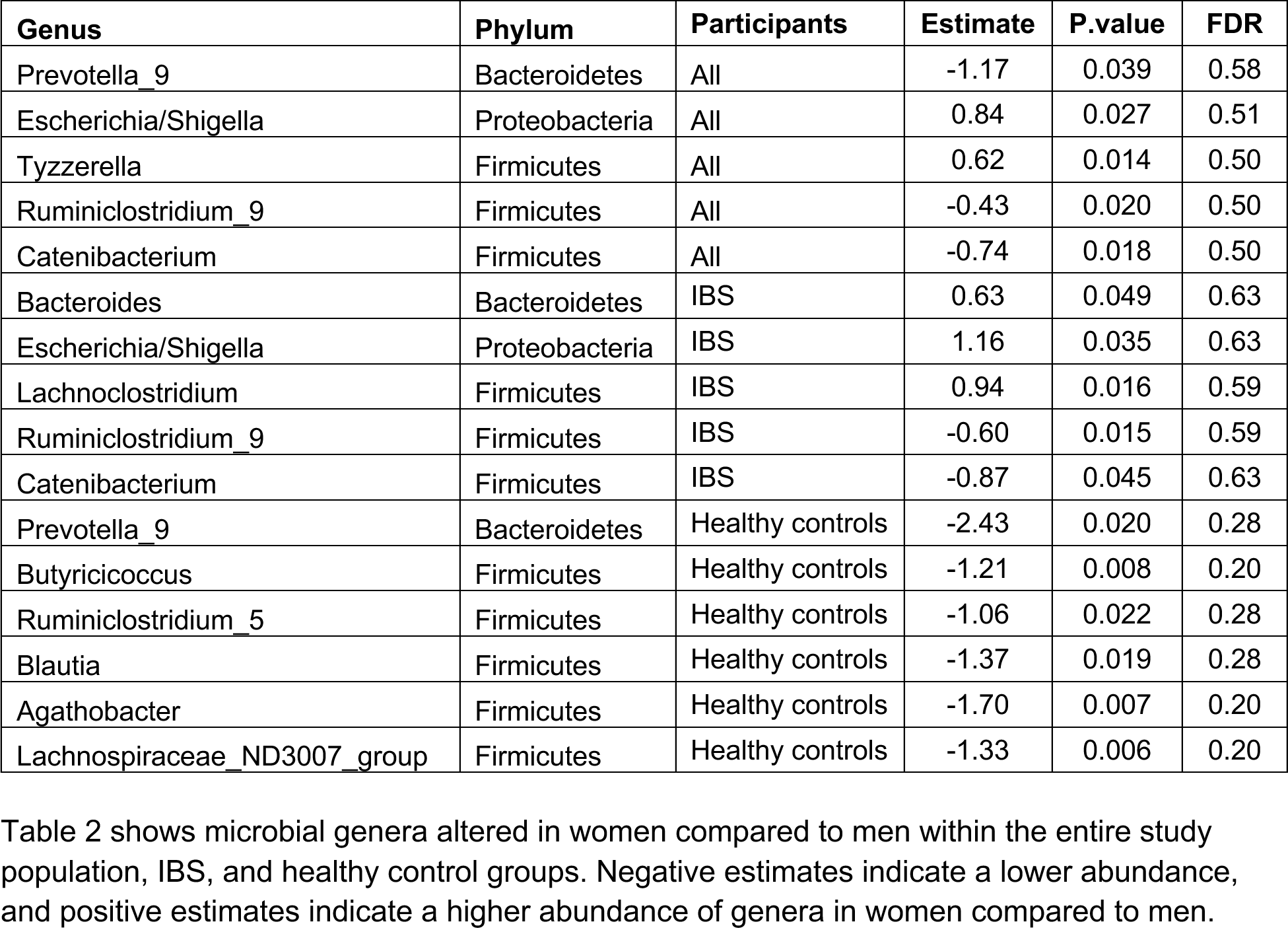
Sex-associated Microbial Genera in IBS Patients and Healthy Controls.

### Sex-related differences in IBS symptoms

Multivariate analysis to evaluate microbial species associated with IBS symptoms identified 20 IBS-symptom-associated genera in men and women. In women, higher symptoms were associated with an increase in abundance of bacterial genera including *Paraprevotella*, *Megasphera*, *Clostridium_sensu_stricto_1*, *Desulphovibrio*, *Candidatus_Arthromonitus* and *Catenibacterium* (**Figure 2A**). In men, however, increased symptoms were associated with an increase of *Staphylococcus*, *Finegoldia*, and *Dialister* (**Figure 2B**). Increased abundances of *Prevotella_7 and Prevotella_9* were associated with higher symptoms in both sexes, but stronger correlations were observed in women. Interestingly, an increased abundance of *Desulfovibrio* was associated with higher symptom severity in women but lower in men.

**Figure 2.**
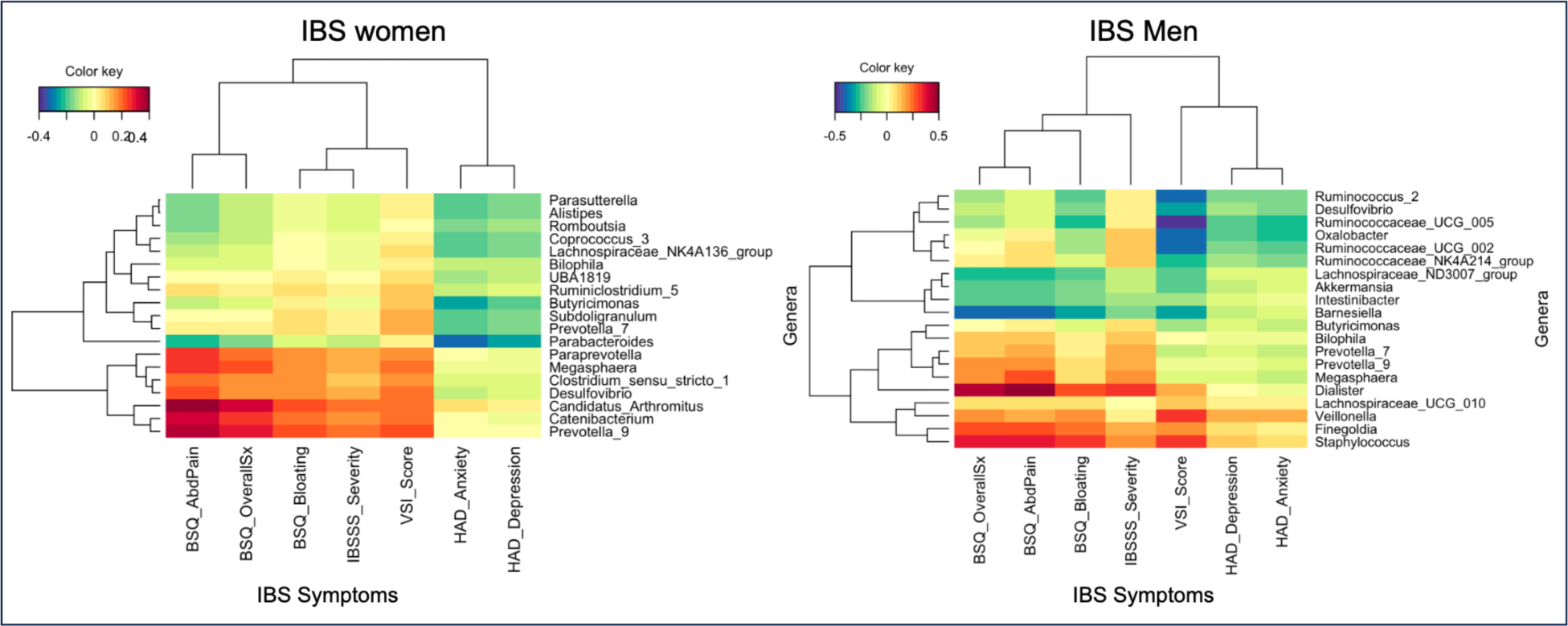
Sex-specific Correlations between Mucosal Microbial Genera and Clinical Symptoms of IBS Patients. The correlation heatmaps show the microbes selected by sparse partial least squared analysis within men and women at genus level that were positively or negatively correlated with IBS symptoms including abdominal pain, bloating, and overall symptom severity. X-axis represents IBS symptom severity measures and Y-axis represents microbes. Red color indicates a positive correlation and blue indicates a negative correlation.

## Discussion

The colonic mucosal microbiome is grossly understudied compared to the fecal microbiome in IBS. Despite growing evidence of sex differences in symptoms and physiologic alterations in IBS, there was previously no data on sex differences in mucosal microbiome in IBS. This is the first study to investigate sex-related differences in mucosal microbiota in IBS and HC. The important findings of our study are that 1) IBS is associated with distinct sex-related differences compared to HC and 2) distinct microbial genera are associated with the severity of IBS symptoms in women compared to men.

The distinct sex differences we identified in the IBS mucosal microbiome compared to HC and their correlation with IBS symptom severity support the importance of these differences in the pathophysiology of IBS. Increased abundance of bacterial genera including *Lachnoclostridium* in IBS women, which is associated with an immune inflamed gene expression profile suggests its potential role in modulating the immune microenvironment contributing to IBS symptoms.^24^ Interestingly, our study identified mucosal-associated *Prevotella* as less abundant in healthy women than healthy men but this was not seen in IBS. Decreased fecal abundance of *Prevotella* in fecal samples has been previously reported in healthy women compared to men. ^25, 26^ In a different and larger cohort of IBS and HC participants, we also found that *Prevotella* was the main driver of sex differences in stool samples of IBS with lower in abundance in women compared to men, but this was not seen in HC. The lower fecal abundance of *Prevotella* was associated with increased IBS symptom severity, while increased mucosal abundance correlated with increased IBS severity in both women and men. Our differing results support prior studies that mucosal and fecal microbiota differ significantly from one another.^27, 28^ Of note, *Prevotella* is a highly heterogeneous genus and some species can be beneficial, while others that can be pathogenic.^29^ Due to the resolution of 16S sequencing, we could not determine which species of *Prevotella* were associated with sex differences in the stool and mucosal samples.

Our study identified other sex-specific associations of IBS symptoms with bacterial genera. Increased *Desulfovibrio* was associated with lower symptoms in men but higher symptoms in women. *Desulphovibrio* is a sulfate-reducing bacteria (SRB) known for its direct association with increased hydrogen sulfide production and high-fat diet-induced colitis.^24^ One potential mechanism linking microbes to sex-specific disease association may be through sex hormones. For example, studies suggest that estrogen affects the expression of toll-like receptors (TLRs), which induce a pro-inflammatory environment that facilitates the translocation of bacteria into the lamina propria thereby amplifying the pro-inflammatory responses.^30^ These sexually dimorphic effects of microbes on IBS symptoms emphasize the necessity for sex-specific investigations into the role of microbiota composition in IBS.

Some of the bacteria we observed as differentially abundant between sexes were also reported in the context of various diseases and health (**Supplementary Table 1**). Similarly, *Clostridium_sensu_stricto_1*, an opportunistic pathogen, which was associated with increased symptoms in women but not men in our study, is also reported to be increased in women with chronic fatigue syndrome (CFS),^31^ which is a known comorbidity of IBS, predominantly women. Another genus, *Catenibacterium*, which was less abundant in the colonic mucosa of IBS women compared to men, was found to be lower in IBS compared to healthy controls. It is a butyrate-producing bacteria known to improve intestinal barrier function,^32^ and has also been reported to be less abundant in women compared to men,^33^ (**Supplementary Table 1**).

There are some limitations to this study. We only sampled the sigmoid colonic mucosa but evidence suggests that there are significant differences in mucosal microbiota along the intestinal tract.^25^ While our sample size is lower than fecal microbiome studies, our study has the largest mucosal microbiome sample in the field so far. Metagenomic sequencing was limited to only the specific DNA regions covered by 16S rRNA sequencing. Using shotgun metagenomic sequencing instead would allow us to identify all genomic DNA in the sample.

In conclusion, our study that sex moderates differences in microbial abundances in colonic mucosal tissue and their relationship to IBS symptom severity. The findings indicate the importance of investigating sex-specific mechanisms in-depth and they may differ in the stool vs intestinal mucosa. Further research is warranted to delineate the specific roles of the microbes identified in the pathophysiology of IBS.

## Data Availability

The 16S sequences and associated metadata used in this study are part of several ongoing projects within our research group. Due to the active nature of these projects and our ongoing efforts in writing and publishing additional papers, we are not able to publicly release the entire dataset at this time. However, we are committed to transparency and scientific collaboration. Therefore, specific data requests can be accommodated on a case-by-case basis. We plan to submit the sequence and associated metadata to an appropriate publicly available archive once all ongoing projects are completed.

## Author contribution

SMJ: design of study, analysis, writing, and editing. SS: writing and editing. JL: analysis and editing. TSD: data acquisition and editing. JPJ: data acquisition and editing. LC: design of study, data acquisition, analysis, writing, and funding. SCORE group: writing/editing, data acquisition.

---

**Supplementary Table 1. IBS and Sex-associated Microbiota Reported in Literature**

Supplementary Table 1 shows a list of studies that investigated microbes that were found to be associated with sex within IBS or healthy controls in this study.

